# Development and evaluation of codelists for identifying marginalised groups in primary care

**DOI:** 10.1101/2024.09.11.24313391

**Authors:** Tetyana Perchyk, Isabella de Vere Hunt, Brian D Nicholson, Luke Mounce, Kate Sykes, Yoryos Lyratzopoulos, Agnieszka Lemanska, Katriina L Whitaker, Robert S Kerrison

## Abstract

**Background:** Primary care electronic health records provide a rich source of information for inequalities research. However, the reliability and validity of the research derived from these records depends on the completeness and resolution of the codelists used to identify marginalised populations.

**Aim:** The aim of this project was to develop comprehensive codelists for identifying ethnic minorities, people with learning disabilities (LD), people with severe mental illness (SMI) and people who are transgender.

**Design and setting:** This study was a codelist development project, conducted using primary care data from the United Kingdom.

**Method:** Groups of interest were defined a priori. Relevant clinical codes were identified by searching Clinical Practice Research Datalink (CPRD) publications, codelist repositories and the CPRD code browser. Relevant codelists were downloaded and merged according to marginalised group. Duplicates were removed and remaining codes reviewed by two general practitioners. Comprehensiveness was assessed in a representative CPRD population of 10,966,759 people, by comparing the frequencies of individuals identified when using the curated codelists, compared to commonly used alternatives.

**Results:** A total of 52 codelists were identified. 1,420 unique codes were selected after removal of duplicates and GP review. Compared with comparator codelists, an additional 48,017 (76.6%), 52,953 (68.9%) and 508 (36.9%) people with a LD, SMI or transgender code were identified. The frequencies identified for ethnicity were consistent with expectations for the UK population.

**Conclusion:** The codelists curated through this project will improve inequalities research by improving standards of identifying marginalised groups in primary care data.

**HOW THIS FITS IN:**

- The reliability and validity of primary care data for inequalities research depends on the comprehensiveness of the codes used to identify people from marginalised groups.
- This study set out to develop comprehensive codelists for the identification of four key groups, known to experience health inequalities.
- We developed comprehensive codelists for identifying ethnic minorities, learning disabilities, severe mental illness and people who are transgender, using a systematic approach.
- The codelists were validated by two general practitioners, assessed in a representative sample, and can now be used in primary care practice and research, both nationally and internationally.

## 1. INTRODUCTION

Health inequalities are unfair and avoidable differences in health, which are systematically established and arise from the social conditions in which people are born, grow, live, work and age [1].

In the United Kingdom (UK), health inequalities are a national priority [2]. The National Health Service for England (NHS England) developed ‘The CORE20Plus5 Framework’, which outlines priority groups and clinical areas requiring accelerated improvement [3]. The ‘Core20’ refers to the 20% most deprived areas in England, while the ‘Plus’ refers to population groups that should be identified at a local level, including (but not limited to): ethnic minority groups, people with a learning disability (LD) or autism, people with multiple long-term health conditions, other groups that share protected characteristics (as defined by the Equality Act 2010 [4]), and groups experiencing social exclusion (such as coastal communities, where there may be small areas of high deprivation hidden amongst relative affluence) [3]. The ‘5’ describes key clinical areas requiring accelerated improvement for these populations, namely: maternity, severe mental illness (SMI), chronic respiratory disease, early cancer diagnosis and hypertension case-finding and management [3].

While the CORE20Plus5 framework provides a useful outline of national priorities for health research and clinical practice, there are several ongoing challenges that need to be addressed for researchers to support national strategy [5]. Pertinent among these is the identification and classification of CORE20Plus groups (also referred to as ‘socially disadvantaged’ or ‘marginalised’ groups) within electronic health records (EHRs) [5]. EHRs include key administrative clinical data relevant to a person’s care, including demographics, progress notes, problems, medications, vital signs, past medical history, immunisations, laboratory data and radiology reports [6]. These data are recorded using clinical coding systems, typically containing hundreds of thousands of concepts [7], which need to be categorised via codelists to facilitate research.

While methods for codelist development and reporting have previously been published [8-10], and many studies now report the methods used to develop codelists, no papers describing the systematic development of codelists for identifying marginalised groups appear to exist [9].

To support future health inequalities research, we set out to develop comprehensive codelists for the identification of four CORE20plus5 groups, namely: people from ethnic minority groups, people with LDs, people with SMI and people who are transgender. We selected these groups, specifically, for two reasons. First, this codelist development project formed part of a wider study exploring inequalities in breast cancer diagnosis and treatment, and these groups are all known to experience inequalities in breast cancer outcomes. Second, the resources required to curate codelists for all CORE20Plus groups were not available.

## 2. METHODS

Previously published approaches for codelist development were reviewed [10], and Watson et al’s approach was deemed the most appropriate because it specifically sets out methods for developing codelists using UK based EHRs.

The approach comprises three steps: **Step 1**. Clearly define the clinical feature of interest a priori; **Step 2**. Assemble a list of codes that may be used to record the clinical feature; **Step 3**. Delphi review of codes.

The following presents a detailed overview of the processes undertaken for each step.

### 2.1. Step 1. Clearly define the clinical feature of interest a priori

Watson et al recommend beginning by clearly defining the clinical feature of interest [11]. To do this, they recommend using reliable sources of information, such as the National Institute for Health and Care Excellence (NICE) [12]. For the purposes of this study, we used the following definitions and sources to define marginalised groups of interest:

#### Ethnicity

The Office of National Statistics (ONS) has highlighted that there is no true consensus on what defines an ethnic group, as identification to these is “self-defined and subjectively meaningful to the individual” [13]. However, as stated by NHS England, it is generally accepted that ethnicity includes a variety of elements, such as ancestry, culture, identity, religion, language, and physical appearance [14].

We defined ethnicity according to the characteristics set out in the Equality Act 2010, namely: colour, nationality and ethnic or national origins [4]. To this end, UK Census (2021) categories were used to group ethnicity codes (see table 1) [15].

**Table 1.** UK census ethnic categories [15].

|  |
| --- |
| **Table 1.** UK census ethnic categories [15]. |
| **Asian or Asian British** |
| Indian |
| Pakistani |
| Bangladeshi |
| Chinese |
| Any other Asian background |
| **Black, Black British, Caribbean or African** |
| Caribbean |
| African |
| Any other Black, Black British, or Caribbean background |
| **Mixed or multiple ethnic groups** |
| White and Black Caribbean |
| White and Black African |
| White and Asian |
| Any other Mixed or multiple ethnic background |
| **White** |
| English, Welsh, Scottish, Northern Irish or British |
| Irish |
| Gypsy or Irish Traveller |
| Roma |
| Any other White background |
| **Other ethnic group** |
| Arab |
| Any other ethnic group |

#### Learning disability

The Department of Health and Social Care (DHSC) defines a learning disability as: “a significantly reduced ability to understand new or complex information, to learn new skills (impaired intelligence), with a reduced ability to cope independently (impaired social functioning), which started before adulthood” [16]. According to Public Health England: “a learning disability is different to a learning difficulty, which is a reduced intellectual ability for a specific form of learning and includes conditions such as dyslexia (reading), dyspraxia (affecting physical co-ordination) and attention deficit hyperactivity disorder (ADHD)” [17].

We defined learning disabilities according to the characteristics described by the DHSC. We did not categorise learning disabilities into groups. They are often described as being ‘mild’, ‘moderate’, or ‘severe’, ‘verbal’ or ‘non-verbal’, and ‘syndromic’ or ‘non-syndromic’, but there is no consensus as to which learning disabilities fall into which categories, and there is individual-level variation among individuals with the same disability, making it inappropriate to generalise [18].

#### Severe mental illness

The Diagnostic and Statistical Manual, fifth edition (DSM-V), defines severe mental illness as: “a mental, behavioural, or emotional disorder resulting in serious functional impairment, which substantially interferes with or limits one or more major life activities.” [19].

We defined SMI according to the characteristics described in the DSM-V, and categorised codes according to four recognised conditions, namely: ‘schizophrenia’, ‘bipolar disorder’, ‘severe major depression’ and ‘other psychotic disorders’ [19].

#### Transgender

The UK Government Equality Office defines trans (transgender) people as “People whose gender is different from the gender assigned to them at birth” [20]. They provide the following by way of example: “A trans man is someone that transitioned from woman to man” [20].

We used the definition provided by the Government Equality Office [20]. Most clinical codes do not differentiate between trans men and trans women; therefore, we did not group codes according to the direction of the transition (i.e. male to female; female to male).

### 2.2. Step 2. Assemble list of codes that may be used to record the clinical feature

In the second step, an online thesaurus was used to develop a list of synonyms associated with the clinical features of interest. An overview of the synonyms used for each clinical feature is presented in Table 2.

**Table 2.** List of synonyms used for each clinical feature.

| **Table 2.** List of synonyms used for each clinical feature |  |
| --- | --- |
| **Clinical feature** | **Synonyms** |
| **Ethnicity** | Nationality, Nationalities, Nation(s), Race(s), Racial, Colour, Ethnic, Ethnicity, Minority, Minorities |
| **Learning Disability** | Intellectual disability, Intellectual disabilities, Learning disability, Learning Disabilities, ID(s), LD(s) |
| **Severe mental illness** | Serious mental illness, Serious mental illnesses, Severe mental illness, Severe mental illnesses, SMI(s) |
| **Transgender** | Trans, Transsexual, Transgender, Transgender, Gender dysmorphia, Trans man, Trans woman, Trans men, Trans women, Transfeminine, Transmasculine, Gender Surgery, LGBT. |

We then searched for published codelists, relating to the clinical features, using the synonyms identified. One major resource used was the October 2023 Clinical Practice Research Datalink (CPRD) Bibliography [21]. CPRD is one of the largest research databases of electronic, anonymised, and longitudinal medical records from primary care in the world [22]. The CPRD Bibliography is a resource which provides a record of all published studies (>3,000 as of October 2023) using CPRD data. The bibliography aided in our search for relevant publications. The codelists used in these publications were subsequently downloaded, where they had been made available.

To capture codelists used in non-CPRD studies, we also searched for, and downloaded, relevant codelists from four widely used codelist repositories, namely: OpenCodelists [23], the Health Data Research UK [HDRUK] Phenotype Library [24], the University of Cambridge Codelist Repository [25] and the London School of Hygiene and Tropical Medicine Data Compass [26].

The codes from the codelists were then extracted and collated into separate excel files for ethnicity, LD, SMI and transgender. Duplicate codes were removed, and remaining codes grouped by the two main coding systems used in the UK: Read and SNOMED-CT. Read was generally used in primary care until 2018, and SNOMED-CT thereafter, following the enablement of SNOMED-CT within Primary care systems [27].

The aggregated codelists were then checked for accuracy using the CPRD code browser, which contains the diagnostic description, the alphanumeric Read / SNOMED-CT code originally used by the general practitioner (GP) to enter the clinical information, and CPRD’s proprietary ‘medcode’ / ‘medcodeid’ (which is simply a numeric equivalent of the Read / SNOMED-CT code) [27]. The tool allowed for easy browsing by either using a known code or description to check code accuracy. In line with the method described by Watson et al [11], inaccurate codes were removed or corrected, and additional codes (not previously identified) added if they were associated with the diagnostic definition of one or more eligible codes.

### 2.3. Step 3. Delphi review of codes

In the final step, the codelists were sent to a GP (IdVH) for review. They scored each code using the following three-point scale:

1. Definitely include: the code accurately defines the clinical feature of interest, and GPs would use it.
2. Uncertain: it remains unclear whether the code accurately reflects the clinical feature of interest, or whether GPs would use it.
3. Definitely exclude: the code does not define the clinical feature of interest, and GPs would avoid.

Codes assigned a score of 2 (‘Uncertain’) or 3 (‘Definitely exclude’) were reviewed by a second GP (BN). All codes assigned a score of 3 (‘Definitely exclude’) by both reviewers were removed from the codelists. All codes receiving a score of 2 from either reviewer were retained as ‘uncertainty variables’, which are recommended for sensitivity analyses [11]. These variables are typically less frequently used by GPs, but may identify additional, potentially relevant, cases [11].

### 2.4. Assessing codelist comprehensiveness

To explore the comprehensiveness of the developed codelists, we used the CPRD database browser to ascertain the number of people with at least one recording of an included code for each list. We restricted the population to those who had been registered with their CPRD Aurum practice since at least 1^st^ January 2019 and who had neither died nor deregistered from their practice by August 2022 (the date of last data collection).

In the context of CPRD studies, researchers have been able to acquire already categorised ethnicity data through linkage with the Hospital Episode Statistic (HES) [28]. Derived ethnicity data is also now available through CPRD at an additional cost [29]. The ethnicity codelist we generated will service to those researchers who are using other non-CPRD datasets, or ones who may not have access to either the linked HES data or the CPRD derived ethnicity.

For the other variables, validated codelists from other sources were selected as comparisons, seeking to match the scope of each list as closely as possible. Codelists for LD and SMI were sourced from the Quality and Outcomes Framework (QOF: the pay for performance scheme for general practices in the UK) [30]. QOF includes incentives for practices to keep a register of patients with certain conditions, including LD and SMI, and produces validated codelists for this purpose. We used lists from the QOF Business Rules version 45; “Learning Disability codes” (list name LD_COD) and “Psychosis and schizophrenia and bipolar affective disease codes” (list MH_COD). For the transgender category, no QOF list exists, and literature is slim. We identified two identical previously published codelists that were used to compare to our newly developed codelist.

## 3. RESULTS

As demonstrated in figure 1, a total of 52 codelists were identified. After removing duplicates and Delphi review, 1,420 unique codes remained.

**Figure 1.**
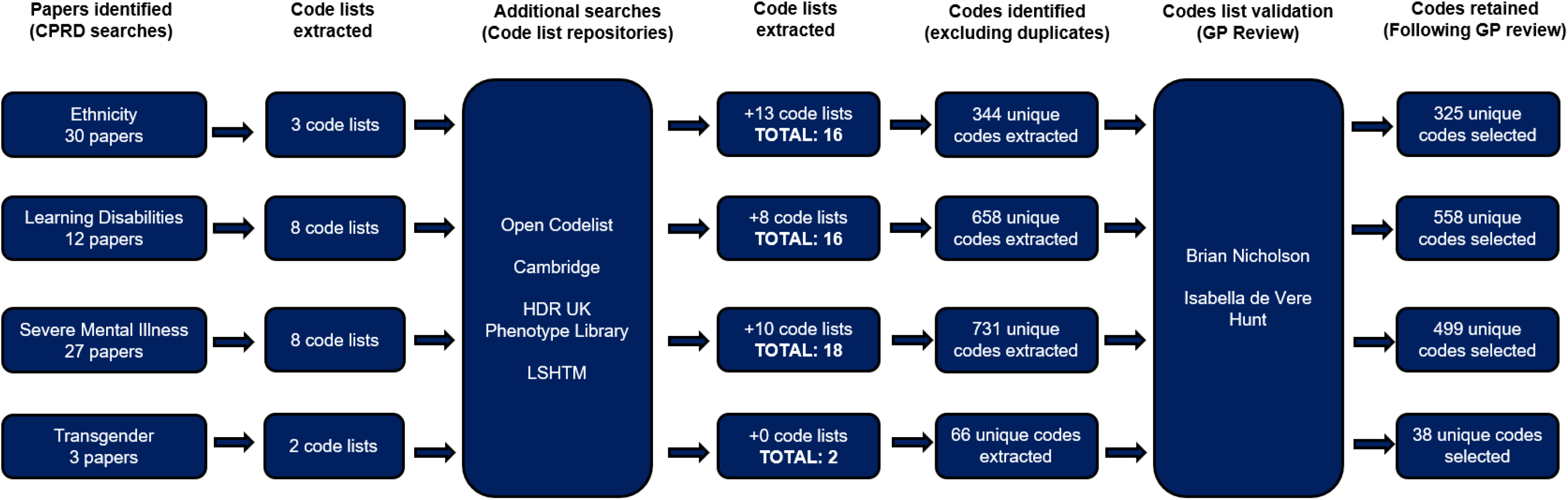
Overview of codelist development.

The codes were run in the source population of CPRD (n=10,966,759) (see Table 3). 110,692 patients had one or more codes for LD (1.00%), 129,788 for SMI (1.18%) and 1,884 for transgender (0.02%). This represented an increase of 48,017 (76.6%) for LD, 52,953 (68.9%) for SMI, and 508 (36.9%) for transgender (see Table 3).

**Table 3.** Comparing frequencies derived from original and curated codelist for LD, SMI and transgender as well as frequencies between ethnic groups.

|  |  |  |  |  |
| --- | --- | --- | --- | --- |
| Total population (N) | Mean age (SD*) | Age range | Frequency of males (%) | Frequency of females (%) |
| 10,966,759 | 43.3 (22.8) | 4 – 100 years old | 5,554,024 (50.6) | 5,412,735 (49.4) |
| Inequality Group | Original Codelist (N) | Curated Codelist (N) | Difference in Identification (N) | Percent Increase (%) |
| Learning disability | 62,675 | 110,692 | 48,017 | 76.6 |
| Severe mental illness | 76,835 | 129,788 | 52,953 | 68.9 |
| Transgender | 1,376 | 1,884 | 508 | 36.9 |
| Ethnicity |  | Frequency from curated codelist (N) |  | Frequency percent (%) |
| White |  | 3,112,319 |  | 71.0 |
| Asian or Asian British |  | 285,126 |  | 6.50 |
| Black, Black British, Caribbean or African |  | 116,570 |  | 2.66 |
| Mixed or multiple ethnic groups |  | 63,024 |  | 1.44 |
| Other ethnic group |  | 94,693 |  | 2.16 |
| Unknown |  | 712,286 |  | 16.3 |
| *SD = Standard deviation |  |  |  |  |

Ethnicity data were available for 4,384,017 (39.98%) individuals. Of these, 3,112,319 (71.0%) were white, 285,126 (6.50%) Asian, 116,570 (2.66%) black, 63,024 (1.44%) mixed, 94,693 (2.16%) other and 712,286 (16.3%) unknown (see table 3).

## 4. DISCUSSION

### 4.1. Summary

We created comprehensive codelists for identifying ethnicity, LD, SMI and transgender records in EHRs. Compared with the commonly used alternatives, our codelists improved data capture by 76.6% for LD, 68.9% for SMI and 36.9% for transgender records. These codelists have been made publicly available for others to use in their own research: https://osf.io/8skze/. They are readily available for use in UK research, but need to be adapted for international research, where different code systems are used. We provide code descriptions, alongside codes, to help facilitate this process.

### 4.2. Strengths and limitations

This study has several strengths. First, code lists were identified through multiple sources, including CPRD publications and code list repositories, maximising the chances of identifying potentially relevant codes. Second, instead of accepting codes at face value, a Delphi review was implemented, with potentially irrelevant codes removed or segregated for sensitivity analysis.

This study also has a number of limitations. First, it only used codes from code lists arising from CPRD publications, or uploaded to code list repositories, and, despite several safety netting procedures, it is possible not all relevant codes were captured. This is exemplified by the fact that only 29% of relevant publications made their code lists publicly available, meaning there is potential for many codes to be missed. Second, while we compared the comprehensiveness of our codelists against commonly used alternatives (e.g. QOF), the comparisons were not like-for-like, due to differences in population definitions. For example, the QOF definition of SMI does not include severe major depression, and these differences may partially explain some of the increase observed in our codelist.

### 4.3. Comparison with existing literature

This research builds on the extant literature by synthesising the efforts of those who have previously curated code lists for the identification of marginalised groups. For example, Boyd et al [31] previously created a code list for identifying people who are transgender, which included a total of eight Read codes. Our list includes an additional 30 Read codes (along with the corresponding SNOMET-CT codes), enabling higher resolution and identification of people who are transgender.

### 4.4. Implications for research and / or practice

The findings of our study have several implications for research. First, we found that only a small proportion of researchers made their codelists publicly available. Efforts are therefore needed to encourage researchers to share their codelists. This will expedite inequalities research and prevent duplication of work. Second, while the percentages of ethnicity found in the population closely mirrored the frequencies reported in the UK census 2021 [15], we found that other marginalised groups were under-represented in the CPRD dataset (1.00%, 1.18% and 0.02% were recorded as having a LD, SMI, or transgender code, respectively, compared with national estimates of 2.16%, 2.00-3.00% and 0.20% [32, 33, 34]. Further research is needed to understand why these frequencies do not match national estimates. Finally, there are many more groups experiencing inequalities, for whom we have not developed codelists, including other LGBT groups and long-term conditions. We encourage others to follow our methods to develop comprehensive codelists for the identification of these groups.

The outputs from our study also have implications for policy. For example, the code lists can be used by clinicians for audits and service improvement projects, to ensure correct identification and classification of individuals, and improve the reliability and validity of findings.

## Data Availability

The codelists developed through this work are available from Open Science Framework.

https://osf.io/8skze/

